# Machine learning detects SARS-CoV-2 and variants rapidly on DNA aptamer metasurfaces

**DOI:** 10.1101/2021.08.07.21261749

**Authors:** Hulya Torun, Buse Bilgin, Muslum Ilgu, Cenk Yanik, Numan Batur, Suleyman Celik, Meric Ozturk, Ozlem Dogan, Onder Ergonul, Ihsan Solaroglu, Fusun Can, Mehmet Cengiz Onbasli

## Abstract

COVID-19 is detected using reverse transcription polymerase chain reaction (RT-PCR) of nasal swabs. A very sensitive and rapid detection technique using easily-collected fluids like saliva must be developed for safe and precise mass testing. Here, we introduce a metasurface platform for direct sensing of COVID-19 from unprocessed saliva. We computationally screen gold metasurfaces out of a pattern space of 2^100^ combinations for strongly-enhanced light-virus interaction with machine learning and use it to investigate the presence and concentration of the SARS-CoV-2. We use machine learning to identify the virus from Raman spectra with 95.2% sensitivity and specificity on 36 PCR positive and 33 negative clinical samples and to distinguish wild-type, alpha, and beta variants. Our results could pave the way for effective, safe and quantitative preventive screening and identification of variants.

## Main

Detecting SARS-CoV-2 and its variants rapidly and accurately is an urgent challenge due to the COVID-19 pandemic. The state-of-the-art detection technique for the virus is reverse transcription polymerase chain reaction (RT-PCR) testing. This test is commonly done using nasopharyngeal (NP) swabs, which might not be safe ^1^ or effective ^2^ if not collected properly. Since this test requires expert medical staff and relies on a limited number of certified test laboratories, turnaround times may extend to multiple days ^3,4^. The sensitivities of commercial virus or antigen tests might also be as low as 20% depending on viral load ^2,5^. Food and Drug Administration data on Emergency Use Authorization SARS-CoV-2 virology tests revealed a wide range of limits of detection (LoD), spanning more than 5 orders of magnitude differences. Since each 10-fold increase in the LoD of a COVID-19 viral diagnostic test is expected to increase the false negative rate by 13% ^6^, developing biosensors with high accuracy and a very low LoD is not only important for research but also critical for identifying asymptomatic cases. An important ratio of the cases was found to be asymptomatic and may spread the virus without being detected or isolated ^7^. New SARS-CoV-2 variants started emerging as a significant public health challenge, especially among the unvaccinated. Rapid detection of variants became critical for timely response against potentially immune-evading mutations ^8^. Hence, a highly sensitive and rapid detection technique based on easily-collected body fluids (saliva) must be introduced for mass screening and variant detection regardless of symptoms.

The vast majority of the current SARS-CoV-2 clinical tests ^9,10^ target the viral RNA using RT-PCR ^11^ or the viral antigens using lateral flow assays ^2^. Emerging methods include aptamer probes ^12-14^, Raman-based sensors ^3,15,16^, electrochemical sensing ^17^, field-effect transistors ^18^, plasmonic antibody testing ^19^, waveguide interferometers ^20^, CRISPR ^21^, photothermal ^22^ or resistive sensing ^23^, matrix-assisted laser desorption/ionization time of flight mass spectrometry (MALDI-TOF) ^24^ and deep learning classification of x-ray tomography scans ^25^. While machine learning-driven molecular screening techniques have significantly accelerated drug discovery and the COVID-19 vaccine development ^26-29^, a similar approach is yet to be applied for developing groundbreaking viral sensing technologies.

We developed and applied a genetic algorithm-based nanostructure screening technique for enhancing the Raman and fluorescent scattering cross-section of a DNA aptamer-bound metasurface by more than a few orders of magnitude and developed machine learning classifier models capable of identifying the SARS-CoV-2-specific Raman peaks from clinical saliva samples. Surface-enhanced Raman spectra (SERS) contain molecular fingerprints down to a single nucleotide sensitivity ^30^. Furthermore, unsupervised clustering of these spectra could help identify the emergence of new variants during mass screening. Our unsupervised clustering model identifies the presence of variants B.1.1.7 (alpha, first detected in the UK), B.1.351 (beta, first detected in South Africa), and the wild-type strains.

Figure 1 shows the architecture and the operational workflow of our SARS-CoV-2 saliva biosensor. The sensor consists of a plasmonic 20 nm-thick gold nanopattern on silicon where gold is functionalized with thiol-modified primary DNA aptamers (Fig. **1a**). Unprocessed clinical saliva or inactivated SARS-CoV-2 samples were mixed with Cy5.5-modified fluorescent secondary DNA aptamers and they were allowed to bind for 15 minutes. Next, the solution was drop cast on the chip (2 µL) and left for drying (Fig. **1b**). After drying, the chips were rinsed with double-distilled water and phosphate buffer saline (PBS). Finally, Raman shift spectra within 548–1620 cm^-1^ were measured after exciting the chips with a 633 nm laser (Fig. **1c**). The spectra obtained were entered as input into multiple machine learning classifier algorithms to identify the presence, concentration, and the variants of the virus (Fig. **1d**). The primary DNA aptamer in Fig. **1a** is a spike-specific sequence obtained with the SELEX technique ^13^. The primary DNA is immobilized on the gold metasurface using a thiol attachment. The secondary DNA aptamer includes a Cy5.5 fluorescent marker which yields stronger emission due to localized optical modes built into the metasurface design through the genetic algorithm **(See Supplementary Information)**. Thus, the aptamer-bound surface and the secondary DNA aptamer confine the virus as a sandwich assay for strong light-matter interaction and emission **(Supplementary Fig. S1)**.

**Fig. 1.**
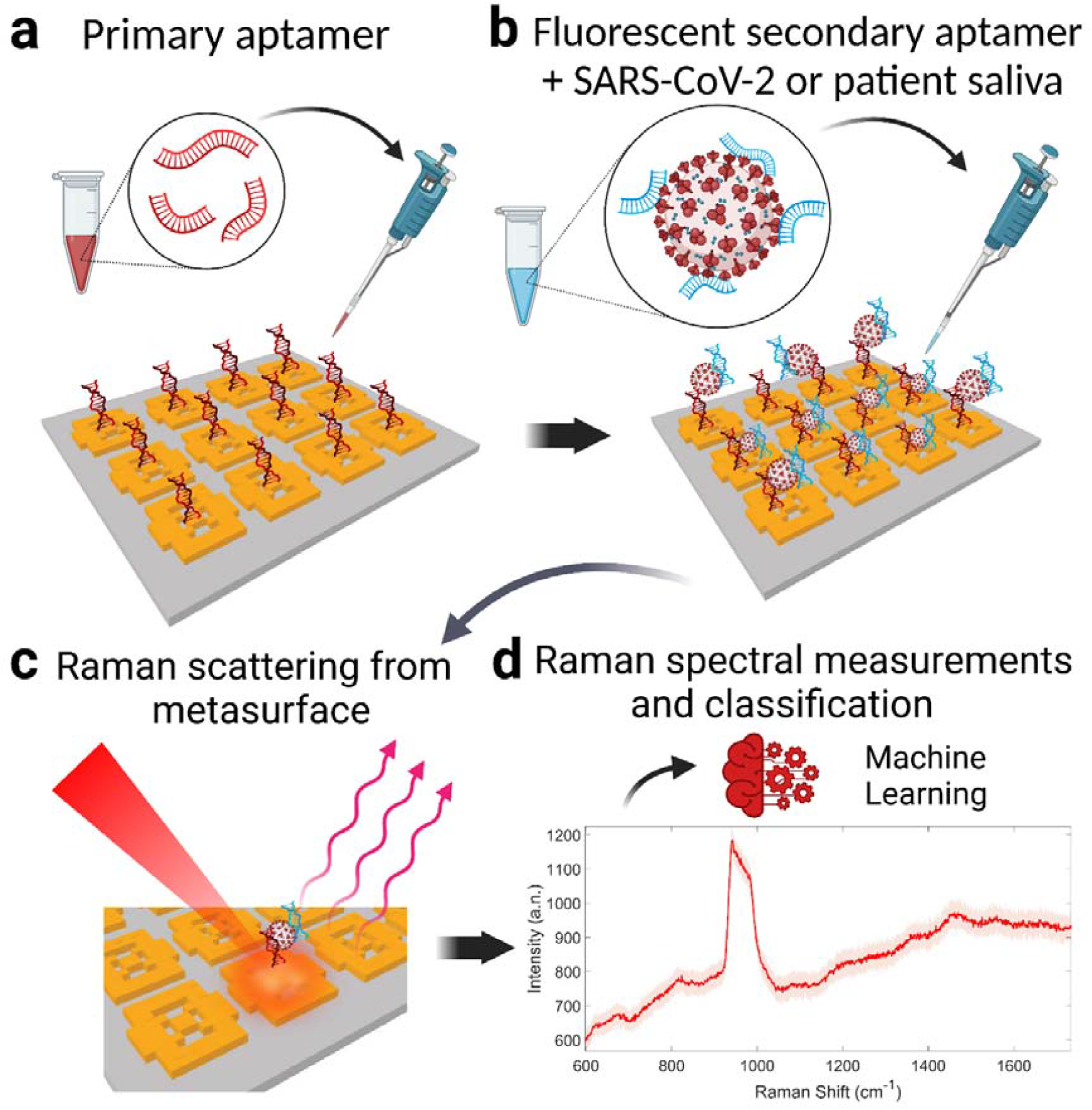
Operation of the SARS-CoV-2 metasurface biosensor with double DNA aptamers and machine learning classification. **a**, The metasurface sensor chip is functionalized using a thiol-modified primary DNA aptamer that is specific for SARS-CoV-2 spike glycoproteins. **b**, Unprocessed COVID-19 patient saliva or inactive SARS-CoV-2 sample is mixed with fluorescent secondary aptamer, resulting in virus-secondary aptamer complex. 2 µL of the mixture is dropped onto the metasurface modified with the primary aptamer to form the sandwich structure (primary aptamer, SARS-CoV-2, secondary aptamer). **c**, Aptamers capture the SARS-CoV-2 by binding to virus glycoproteins, form a sandwich structure yielding strong plasmonic enhancement of the virus-specific fluorescent and Raman emissions. **d**, These Raman spectra are then used in the machine learning classifier model for viral presence, concentration, and the variant type.

## Results

### Development of SERS metasurfaces

A nanostructured metasurface pattern could enhance Raman scattering and help identify a wide dynamic range of viral concentrations. **Fig. 2** shows the high-throughput screening technique for finding the nanostructured metasurface geometries for enhancing the average electric field and fluorescence intensities by at least two orders of magnitude. In our computational screening, we start with an 80-nm square unit cell with 10 × 10 = 100 binary-coded subpixels (1 for gold, 0 for air). We optimize the unit cell for the strongest Raman and fluorescent emission cross-section enhancement while keeping the features large enough for fabrication **(Supplementary Fig. S2)**. A brute force calculation of 3D electromagnetic fields and the fitness functions for all 2^100^ possible unit cells would take 2^100^ × 3 hours/simulation ∼ 4.3×10^26^ years. Running exhaustive parametric screening would take much longer than the time since the Big Bang (13.7 billion years). Thus, a more efficient approach must be used for the high-throughput screening of metasurfaces for sensing. Hence, we used our genetic algorithm to significantly accelerate our screening process.

**Fig. 2.**
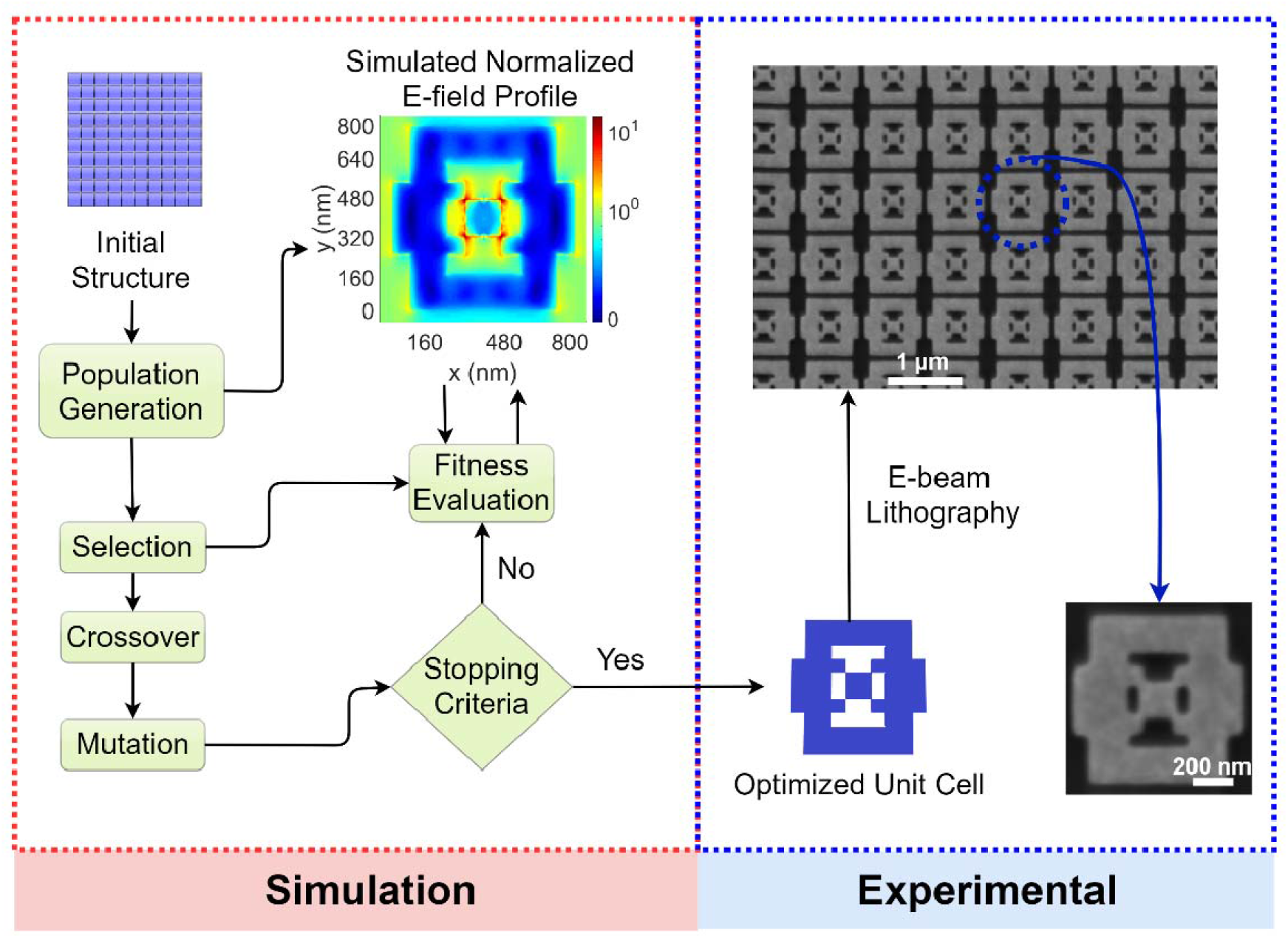
Genetic algorithm-driven computational screening and fabrication of the nanoplasmonic surface-enhanced Raman spectroscopy (SERS) biosensor chip. The genetic algorithm and the flowchart for computational screening of periodic nanostructures for maximizing the Raman cross-section of the metasurface are shown. The electron beam lithography fabrication for the optimized periodic gold nanopattern over 200×200 µm^2^ is shown.

### Sensor characterization and limit of detection

Figure 3 shows the sensor’s detection characteristics. Raman spectra measured for the primary aptamer bound to the chip surface, primary aptamer after adding 2 µl of SARS-CoV-2 virus at 10^8^ pfu/ml concentration, and the sandwich assay, which consists of primary aptamer, SARS-CoV-2, and the fluorescent secondary DNA aptamer (**Fig. 3a**). While virus binding suppresses some of the Raman emission from the aptamer, the fluorescent secondary aptamer amplifies multiple peaks over the entire band of interest (Raman shifts within 600-1700 cm^-1^; 658-710 nm). The Raman spectra of the sensors loaded with 10^2^-10^8^ pfu/ml show a monotonic increase in the emitted peak intensities with higher viral concentrations (**Fig. 3b)**. The peak centered at 970 cm^-1^ (within 920-1020 cm^-1^) is attributed to the silicon substrate’s strong transverse optical phonon line^31,32^ **(Supplementary Fig. S3)**. This peak does not overlap with the higher Raman shift peaks, which are essential for identifying the virus.

**Fig. 3.**
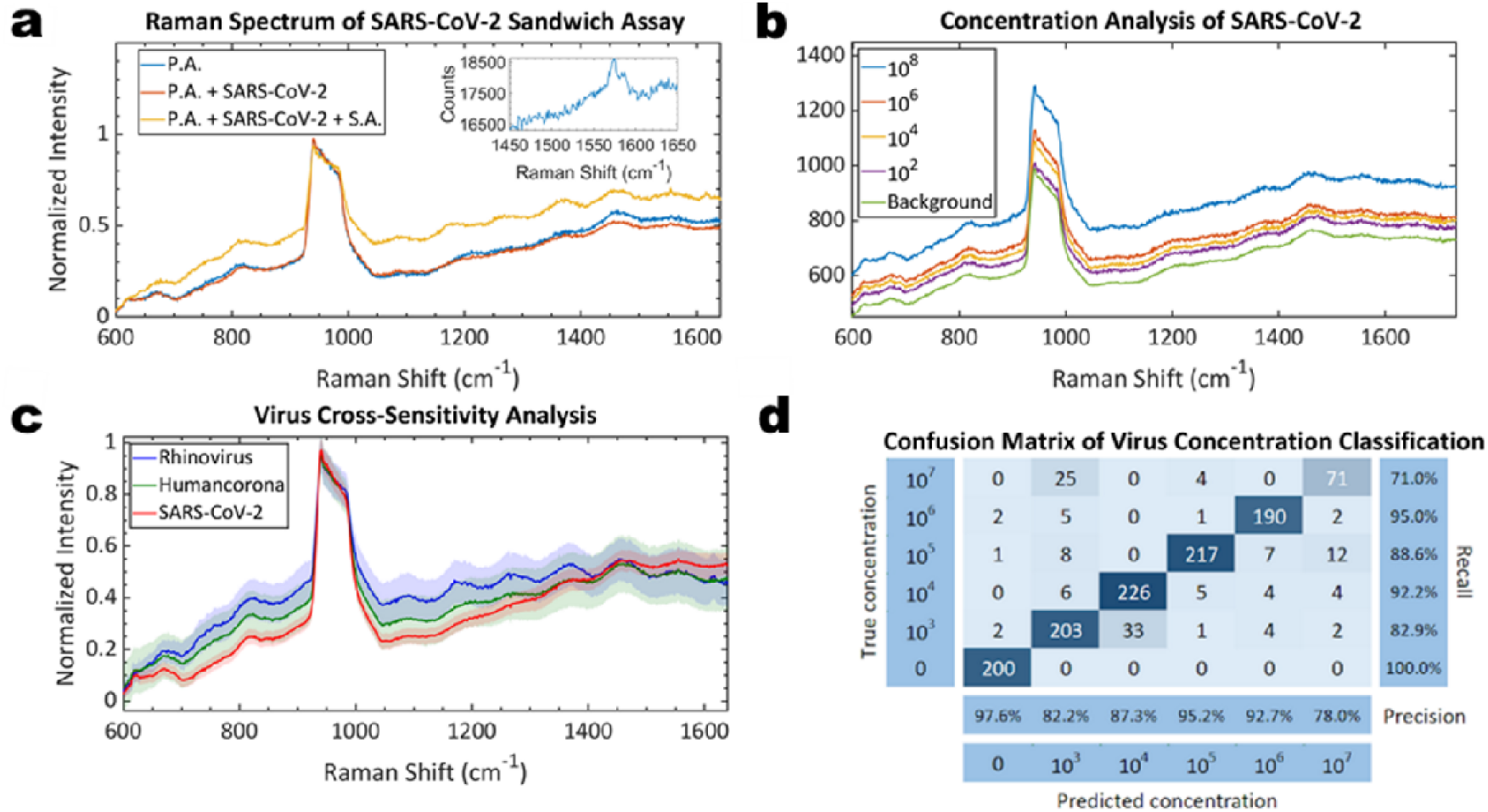
Sensor detection characteristics, cross-sensitivity, and spectrum classification. **a**, The Raman spectra and fluorescence measured for the inactivated aptamer-virus sandwich assay yields distinct features which help identify the presence of the virus (inset: Pure isolated SARS-CoV-2 virus with 10^8^ pfu/ml on CaF_2_ substrate shows the peak near 1575 cm^-1^ attributed to the virus). **b**, Increasing the concentrations of inactivated SARS-CoV-2 (10^2^-10^8^ pfu/ml) manifests clear distinctions in the Raman spectra, fluorescent slope, and emission strength. **c**, Normalized Raman spectra and the error ranges for the rhinovirus, human coronavirus, and inactivated SARS-CoV-2 show the background and peak contrasts, indicating the distinctions in Raman emission characteristics. **d**, The confusion matrix for the machine learning classification of the Raman spectra obtained for each different chip with different inactive viral concentrations (10^3^ to 10^7^ pfu/ml) shows the high detection accuracy of the model based on the Raman spectral features.

### Cross-sensitivity tests

To test cross-sensitivity, we loaded our chips with rhinovirus, human coronavirus, and SARS-CoV-2 and found highly different Raman emissions in the normalized spectra, suggesting the aptamer binding to only SARS-CoV-2 (**Fig. 3c**). A support vector machine (SVM) classification model was trained and tested using these spectra and could identify and distinguish these viruses with 99.7% cumulative variance **(Supplementary Fig. S4)**.

**Fig. 3d** shows the confusion matrix output for the machine learning model that we trained to classify a total of 1235 spectra measured for 10^3^ to 10^7^ pfu/ml concentrations. For each concentration, Raman spectra were measured from 1000 different locations on the chips and our nonlinear machine learning model accounts for non-monotonic or other complex peak intensity changes with concentration and wavelength.

### Clinical trial results

The sensor characterization in Fig. 3 yields about 99% sensitivity and specificity for the multi-class classification of the viral concentrations and the presence of the virus. The clinical validation of the sensor (**Fig. 4**) using unprocessed saliva also yields a similar sensitivity and specificity of 95.2% each. **Fig. 4a** shows the histogram of the PCR cycle threshold (CT) values of the saliva samples, indicating a clinical viral load range of 10^3^-10^7^ pfu/ml ^33^. While saliva maintains a neutral pH within 6.2-7.5, the composition may significantly vary among clinical samples and yield a complex Raman background. As a result, we use a support vector machine and linear discriminant analysis for classifying the spectra as COVID-19 positive or negative. The confusion matrix of our model, **Fig. 4b**, shows only 1 false positive and 1 false negative with a 95.2% sensitivity and 95.2% specificity out of 69 clinical samples in a variety of training/testing configurations **(Supplementary Fig. S5, S6)**.

**Fig. 4.**
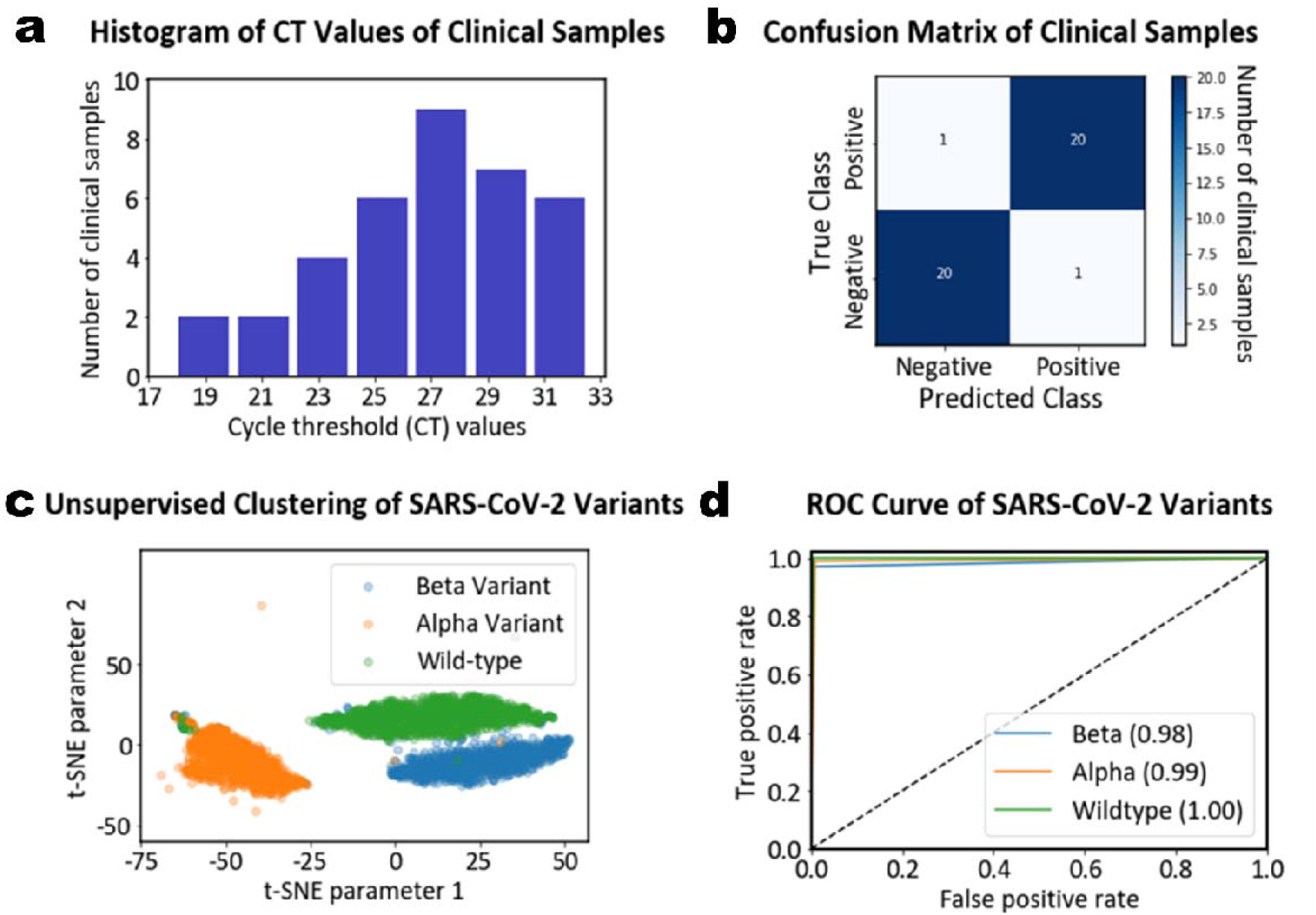
Clinical trial results and variant detection with the sensor. **a**, The histogram of the PCR cycle threshold (CT) values of the clinical samples shows a viral load range of 10^3^-10^7^ pfu/ml used in testing our COVID-19 sensor. **b**, Optimized classification model yields a confusion matrix with only one false positive and one false negative case, yielding 95.2% sensitivity and 95.2% specificity. **c**, Unsupervised clustering model segregates the Raman spectra of inactivated Alpha (B.1.1.7), Beta (B.1.351), and wild-type variants. **d**, Receiver operating characteristics curves show that the unsupervised clustering algorithm distinguishes the variants. The legend shows the area under curve (AUC) values.

### Variant classification results

We developed an unsupervised clustering method for grouping Alpha (B.1.1.7) and Beta (B.1.351) variant samples as shown in **Fig. 4c**. The clustering results indicate that the Raman spectral features might help distinguish the samples as different variants of the virus without next-generation sequencing. A support vector machine model was trained and tested to yield near-perfect accuracy (99.7% cumulative variance) in variant detection **(Supplementary Fig. S4)**.

## Discussion

This study presents a versatile and rapid viral detection technique for SARS-CoV-2 from unprocessed saliva, which might be useful in eliminating the need for collecting nasopharyngeal swabs and in increasing the negative predictive value of the viral tests. In this work; the presence, viral concentration and the variant type of SARS-CoV-2 could be identified from the machine learning classification of Raman spectra of the primary DNA aptamer, the virus and the fluorescent secondary DNA aptamer sandwich assay. Since SERS helps identify the mutations at even a single nucleotide level ^30^, the technique is fundamentally capable of identifying the variants of the virus such as Alpha, Beta (and potentially Delta) by using machine learning classifier models. Hence, the techniques presented here could be used for detecting the emergence of new variants to complement national-wide or global variant screening efforts through whole genome sequencing ^34^. Machine learning also helps eliminate mislabeling of the Raman signals from other viral infections such as rhinovirus or human coronavirus. The Raman spectral features obtained for each virus type is different due to the cross-reactivity of these viruses with the aptamer sandwich assays. Thus, SARS-CoV-2 could be detected and distinguished effectively.

While the focus is on SARS-CoV-2, Raman-based viral detection and machine learning classification technique presented here could be applied for a variety of different viruses and other pathogens. By regularly updating the machine learning classifier models for different viruses or SARS-CoV-2 variants, the detection platform and technique might be used for universal identification of different coronaviruses. Although the DNA aptamers are optimized for SARS-CoV-2 ^13^, the Raman spectral features obtained for each variant in this study have been distinct enough for identification without aptamer replacement. Aptamers can be optimized for any target virus using the SELEX technique and the detection chips could be functionalized with the new DNA or RNA optimal aptamers. Hence, viral detection via machine learning could become mostly a software or machine learning model update and consumable chips might also be updated with new optimized aptamers.

For clinical and point-of-care adoption of the Raman-based detection system presented here, a few key challenges might need to be addressed. First, the detection optics might need to be miniaturized and automated for practical point-of-care testing in pharmacies or other places with high population densities. Miniaturized Raman spectroscopy systems have been demonstrated to have excellent or lab-equivalent performance ^35^ and optimizing these systems for SARS-CoV-2 detection might be feasible. For mass screening purposes, multi-sample trays could be introduced and a digital micromirror device ^36^ or a laser scanner ^37^ might be used for rapid Raman probing and identification of the virus among many clinical samples.

Second important challenge is saliva sample processing. Since the oral bacterial flora could contain more than 700 bacterial species or phylotypes ^38^, saliva sample compositions and their respective Raman spectra might vary dramatically among different people with different oral hygiene practices. In this case, introducing oral rinse steps before saliva sample collection could help reduce the Raman background that could originate from a diverse oral bacterial flora. In addition, Raman spectrum acquisition from multiple points over the saliva samples and additional image and Raman spectrum filtering could help eliminate contaminated background. Thus, the detection accuracies in larger clinical cohorts might be enhanced by introducing a combined methodology of sample preprocessing and spectrum post-processing.

Overall, infrared spectroscopy of biomolecules on photonic metasurfaces aided by machine or deep learning has been a highly promising area with key breakthroughs for sensing, including detection of all major biomolecules ^39^, protein fingerprinting ^40^, early sepsis detection ^41,42^. By adding SERS, we significantly broaden the palette of applications available for medical diagnostics.

## Conclusions

Our results show that COVID-19 and the emergence of its variants can be detected rapidly with high sensitivity and specificity using machine learning and SERS metasurfaces functionalized with DNA aptamers. The ability to optimize for the highest figures of merit over any multi-objective photonic functionality using the genetic algorithm screening presented here also enables new possibilities for massively accelerating materials and device geometry screening for new sensor development. The machine learning and unsupervised clustering models presented here could help identify emerging variants or off-seasonal increases ^43^ of other respiratory viruses during population-scale tests ^8^.

## Methods

### SARS-CoV-2 Isolation from Clinical Samples

Nasopharyngeal and oropharyngeal swab and saliva samples from SARS-CoV-2 polymerase chain reaction (PCR) positive patients were collected with synthetic swabs on the third day after diagnosis. Each swab was placed in a separate sterile tube containing 3 ml of viral transport medium and sent to the Biosafety Level 3 (BSL-3) Laboratory at Koç University Hospital for virus isolation. In the BSL-3 laboratory, samples were aliquoted in 1 ml volume and frozen at -80°C until the virus isolation and all the virus isolation studies were performed in BSL-3 laboratories.

Vero CCL-81 cells grown in Dulbecco minimal essential medium (DMEM) supplemented with antibiotic/antimycotic (GIBCO) and heat-inactivated fetal bovine serum (5% or 10%) were used for SARS-CoV-2 isolation and first passage. Vero cells were adjusted to 2.5 × 10^5^ cells/ml in DMEM containing 10% FBS and antibiotic/antimycotic. Serial dilutions of the clinical nasopharynx and oropharynx samples were prepared in 96-well plates at 100 μl volume with DMEM. Then, 100 μl of Vero cell suspension was added on to serial dilutions of the clinical samples. Inoculated cultures were monitored for cytopathic effect (CPE) daily for 5 days at 37°C and 5% CO_2_. At the end of the five-day incubation, the virus titer in the clinical sample was determined as the highest dilution in which the cytopathic effect was inhibited. TCID50 (tissue culture inhibition dose) values were determined by the Carber Method. After the observation of the cytopathic effect, monolayer cells were scraped with a pipette tip and 5 µl of viral lysate was used for nucleic acid isolation for sequencing and reverse transcription PCR (RT-PCR) studies.

### Virus inactivation protocol and validation of virus inactivation

For virus inactivation, β-propiolactone (Sigma-Aldrich, USA) was used ^44^. For this purpose, cell cultures containing the virus were treated with β-propiolactone (1:1000 v:v) at 4^°^C for 24 hours, and the cultures were kept at room temperature for one more day to remove β-propiolactone residues. Virus inactivation was confirmed by the absence of cytopathic effect in two consecutive passages in the VeroE6 cell lines and the inability to demonstrate amplification by quantitative RT-PCR. For quantitative RT-PCR, Center for Disease Control (CDC) test protocols and probes that amplify the N1, N2, and RdRp regions of the virus genome were used ^45^.

### Whole Genome Sequencing (WGS)

Genome sequencing was performed by using Illumina MiSeq system with the Burrows-Wheeler Aligner MEM algorithm (BWAMEM) 0.7.5a-r405 method. Whole genome was amplified by using primers specific to the open reading frame (ORF1b) and N region of SARS-CoV-2. The whole-genome sequence was obtained with the alignment of the overlapping PCR products. The amplicon size for the ORF1b gene region is 132 bp, and 110 bp for the N gene region and the new genome sequence was obtained by matching with the SARS-CoV-2 reference genomes and variant virus genomes. For aptamer design together with the wild-type virus, Alpha (B.1.1.7) and Beta (B.1.351) variants were provided as well. Poor-quality bases found in the raw data were removed using Trimmomatic 0.36 (-phred33, LEADING:20, TRAILING:20, SLID INGWINDOW:4:20, MINLEN:40). FastQC 0.11.8 was used to evaluate the sequence quality to prevent errors that may occur before cleaning and after the alignment.

### Aptamer modification and design

Aptamers are the short oligonucleotide sequences used in our SARS-CoV-2 biosensor for ensuring that the virus is detected specifically and with high fluorescent amplitude. **Fig. S1** shows the parts used: The primary DNA aptamer **(Fig. S1a)** ^13^ is an oligonucleotide with high affinity (*low nM*) to bind on the receptor-binding domain of SARS-CoV-2 spike glycoproteins. This aptamer has a thiol termination **(Fig. S1c)** for binding to the gold metasurface. The secondary DNA aptamer is modified with Cy5.5, a fluorescent marker **(Fig. S1b)**.

**Fig. S2** shows our experimental confirmation of the virus binding of CoV-2-RBD-4C DNA aptamer ^13^. This aptamer has been modified using Cy5.5 fluorescent marker to be used as the secondary aptamer in this sensor study.

### SARS-CoV-2 Binding Assay

0.5 μM of 4RBD-4C was incubated at room temperature for 10 minutes with only DMEM, or 2×10^4^ or 1×10^5^ of virus particles stored in DMEM without FBS. After collecting bound 4RBD-4C on nitrocellulose filter with vacuum, DNA is extracted with 1M urea and precipitated with 0.3M Sodium acetate and 50% isopropanol. The precipitate was resuspended in RNase free water and PCR with 13 cycles (Denaturation: at 93°C for 3 minutes; Annealing: 93°C for 30 seconds, 65°C for 1 minute, 72°C for 1 minute; Final Extension: 72°C for 10 min) was carried out. Finally, PCR products were separated on 2% agarose gel in 1X TAE Buffer.

Forward and reverse primer sequences are given as follows:

***Oligo3908:*** ATC CAG AGT GAC GCA GCA

***Oligo3909:*** ACG TGT CCA TAT CCG CAA T

### SARS-CoV-2 characterization using Raman spectroscopy

A 5µl of inactivated SARS-CoV-2 virus was dropped onto a CaF_2_ slide (Crystran, CAFP76-26-1U) and the Raman spectrum of inactivated SARS-CoV-2 was recorded applying 50% laser power and 10 seconds of exposure, during 1 Raman scan. The peak around 1575 cm^-1^ in Fig. 3A’s inset is attributed to the virus since there is no other possible peak source in that sample.

### Nanoplasmonic chip design using genetic algorithms and electromagnetic modeling

A nanostructured metasurface pattern could enhance Raman scattering and help identify a wide dynamic range of viral concentrations. To calculate the optimal metasurface geometry that yields the highest average electric field intensity enhancement in **Fig. 2**, we developed a genetic algorithm that generates and “evolves” metasurface geometries and used finite-difference time-domain (FDTD) modeling iteratively to refine the metasurface for the highest figure of merit. We defined the figure of merit or the fitness value η as the average electric field enhancement over a unit cell of the metasurface:

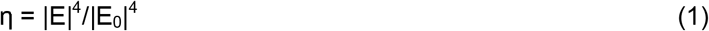

where E_0_ is the total incident electric field and E is the total local electric field summed over all of the meshes in the simulated unit cell. The average field enhancement is a measure for increasing the fluorescence and Raman cross-section of the aptamers and the virus on our chips.

Lumerical FDTD and MATLAB were used for implementing and running the electromagnetic and genetic algorithm models, respectively. In our computational nanostructure screening technique **(Fig. 2)**, we started with an 80 nm-long square unit cell and divided it into a grid of 10 × 10 subpixels (the pixel size and number is limited by electron beam lithography resolution). These subpixels are binary encoded (1 for gold, 0 for air), but they could be numbered differently to include multi-material optimization. The unit cell geometry must be refined to yield the strongest electric field, Raman and fluorescent cross-section enhancement while remaining large enough for fabrication.

The genetic algorithm shown in **Fig. 2** randomly generates a number of binary sequences (population of synthetic nanostructures) and the electric field profiles are calculated for each corresponding nanostructure. The “fitness” values of the structures are calculated, which are the average electric field enhancement values across each unit cell. The nanostructures with the top 20% fitness scores descend to the next “generation” of nanostructures. In preparing the next generation, the structures are mixed and matched with a crossover step and mutations are added as random single or few bit variabilities over the binary sequences. The electric field profiles for the new generation of nanostructures are calculated again and their fitness scores are obtained. The refining procedure is iterated for 30 generations when the enhancement no longer increases. **Fig. S3** shows the evolution of the fitness value stops increasing after 13 generations. The resulting geometry is fabricated as a 20 nm thick gold periodic metasurface pattern using electron beam lithography and lift-off, as shown in **Fig. 2**.

### Nanofabrication of sensor chips

The nanoplasmonic sensor chips were fabricated in Sabancı University Nanotechnology Research and Application Center (SUNUM) Cleanrooms. 4’’ Si wafers were spin-coated with 950 Poly (methyl methacrylate) (PMMA) A4 at 4000 rpm (rotation per minute). The resist thickness is ∼120 nm. Next, the wafer was exposed with Raith EBPG5000 plusES 100 kV electron beam lithography system with a low/small spot size current (∼ 100 pA) and high-resolution parameters at 625 µC cm^-2^ e-beam dose included proximity effect correction (PEC) e-beam module (BEAMER, GenISys GmbH). After exposure, the wafer was developed in 1:3 (by volume) MIBK: IPA (MIBK: methyl isobutyl ketone; IPA: isopropanol) for 1 minute and 1:1 (by volume) concentration MIBK: IPA for 10 seconds, respectively. The wafer was then dipped into IPA for 30 seconds to stop the development, rinsed with IPA, and blow-dried with nitrogen. To eliminate any PMMA resist residues in the exposed areas after development, 7 seconds of oxygen plasma was performed at 50 Watt, 20 sccm O_2_ flow rate, and 37.5 mTorr chamber pressure. After development and plasma cleaning, 2 nm Cr / 20 nm Au layers were thermally evaporated on the wafer. The wafer was dipped in acetone overnight for lift-off. The chips were ultrasonicated in a bath for a short time, rinsed with acetone, isopropanol respectively then blow-dried with nitrogen. Last, the 4” wafer was diced into 5×5 mm^2^ samples where each piece contains a 200 × 200 µm^2^ nanoplasmonic patterned area at the center.

### Surface functionalization of the biosensor chips using oligonucleotides and RNA aptamers and validation using Raman spectroscopy

5’ Thiol C6 S-S modified primary aptamer 3897 (IDT DNA Technologies, 290169003) was activated by incubating with previously prepared 20 µM TCEP (BP Biotechnologies) solution in 1:1 ratio. Aptamer+TCEP solution was diluted with 1X DPBS (BIOWEST, L0615-500) and added to the SERS surface (∼2 µl) with 1 µM of final concentration. SERS substrate was kept under laminar flow for an hour and was washed to remove unbounded aptamers from the surface. SERS substrate was kept in the laminar hood for 15 minutes until it dries. 1M 6-Mercapto 1-hexanol (Sigma-Aldrich, 725226) was added (∼5 µl) to the SERS surface to prevent non-specific binding and SERS substrate was kept under the laminar flow for an hour. The surface was washed with 70% ethanol and rinsed with ddH_2_O.

The Raman spectra of aptamer 3897 were recorded **(Fig. S4)** using a previously calibrated Renishaw InVia™ Raman Microscope equipped with 633 nm excitation wavelength, and 50x objective (Leica 50x/0.75). 10 seconds of laser exposure and 50% laser power (max. of ∼1 mW·μm^-2^ power density) were applied during 1 Raman scan. Cosmic ray removal was performed using Wire 4.4 Software.

Following Raman spectra recording, 5’ Cy5.5™ modified secondary aptamer 1 µM of 3898 (IDT DNA Technologies, 290169004) was added to SERS surface and incubated for binding with primary aptamer 3897 for an hour under laminar flow. SERS surface was washed with 1X DPBS and rinsed with ddH_2_O. The Raman spectra of aptamer 3897 linked 3898 were recorded (Fig. S4) in previously described settings. In **Fig. S4a** contains the background Raman spectrum measurements on silicon, planar gold, and SERS metasurface substrates. The primary and secondary aptamers and their Raman spectra are shown in **Fig. S4b**.

### Nanoplasmonic sensor chip testing under different concentrations of the inactive virus using Raman spectroscopy

Inactive SARS-CoV-2 virus solution with a concentration of ∼1.5×10^8^ pfu/ml was incubated with 1 µM secondary aptamer 3898 for 15 minutes. The stock virus solution was serially diluted to 10^2^ pfu/ml concentration **(Fig. 3b)**. Each SARS-CoV-2 sample (within the concentration range of 10^2^-10^8^ pfu/ml, which were incubated with the secondary aptamer 3898) was added on the primary aptamer bonded SERS surface for 15 minutes. The SERS substrates with different concentrations of SARS-CoV-2 were left to dry for 15 minutes. SERS surfaces were washed with DPBS and rinsed with ddH_2_O. Upon removal of unbound particles from the surface, Raman spectra of SARS-CoV-2 were recorded for each concentration.

### Clinical sample collection and screening

Nasopharyngeal and saliva samples were collected from the patients at Koc University Hospital by Koç University Clinical Trial Unit according to the Koç University Institutional Review Board approval number 2020.112.IRB1.023. Nasopharyngeal samples were tested with PCR as a part of the clinical routine. Saliva samples of the nasopharyngeal COVID-19 PCR positive patients were also tested with PCR and both saliva and the nasal swabs of those patients were ensured to be COVID-19 PCR positive. For 36 patients, both saliva and nasopharyngeal swabs tested COVID-19 positive in PCR. Those 36 PCR positive samples were used in this study to test their saliva with our biosensor. As a negative control, saliva samples of 33 PCR negative healthy voluntary people were collected. Their saliva samples were tested with PCR and have been verified to be COVID-19 negative.

### Clinical validation of the functionalized nanoplasmonic sensor chips

All samples were given to the research team for biosensor measurements by the clinical trials unit in a blunted form of PCR result. Saliva from 69 samples incubated with secondary aptamer 3898 within the ratio of 1:1 in a virus hood in the BSL-2 laboratory. Upon incubation, 2µl of saliva+secondary aptamer 3898 mixture was added onto previously activated primary aptamer 3897 bonded SERS metasurface for 15 minutes. Then, the SERS surface was washed with DPBS and rinsed with ddH_2_O. After wash, Raman spectra of SERS substrates with patient samples were recorded by applying previously described measurement settings.

### Machine learning classifier for characterization of the sensor’s limit of detection

The dataset is gathered by collecting Raman spectra from 6 different samples, each having different virus concentrations. There are 1000 spectra from the virus-free sample, 1225 spectra from each sample with virus concentrations of 10^3^, 10^4^ and 10^5^ pfu/ml, 1000 spectra from the sample with 10^6^ pfu/ml concentration and 500 spectra from the sample with 10^7^ pfu/ml concentration **(Fig. 3d)**.

%80 of the collected dataset is used for training and the remaining 20% is used for tests. A variety of preprocessing pipelines is applied to the data and the classification accuracies of these pipelines are compared with each other. Applying spectra-wise normalization and Principal Component Analysis (explaining 90% of the data with 127 components) yielded the highest accuracy results, hence were chosen to be the preprocessing method. After that, a Support Vector Machine (SVM) model with a linear kernel is trained on the training set with 5-fold cross-validation. The optimal “C” value of the SVM is determined as 0.1, and the decision function shape is chosen to be “One versus One (OVO)”. The training accuracy is recorded as 88.76%. The confusion matrix regarding the test results of the model is given in **Figure 3d**.

### Machine learning classifier for cross-sensitivity

We used different metasurface chips for SARS-CoV-2, human rhinovirus 1B (ATCC® VR-1645™), and human coronavirus 229E (ATCC® VR-740™) to measure their Raman spectra on 425 or 500 different points from each chip. Each spectrum contains 1020 data points. For each virus (SARS-CoV-2, rhino, human CoV); 425, 425 and 500 spectra were measured, respectively. 80% of the dataset was used for training while 20% was used for testing. 5-fold cross-validation was used. A support vector machine with a linear kernel was used with C = 0.1 **(Fig. 3c and Fig.S5)**

To identify the virus type, we used feature scaling and principal component analysis. Only two principal components are sufficient to yield 99.7% cumulative explained variance. Similar results are obtained when the preprocessing method is changed from feature scaling to sample-wise normalization and hence, the classification method is robust under the given measurement conditions. **Fig. S5** shows the results for the cross-sensitivity analysis. **Fig. S5a** shows the principal components I and II and how the spectra are distributed based on this analysis. **Fig. S5b** shows the confusion matrix for this model, where the horizontal axis shows the predicted label and the vertical axis is for the true label. The figure shows that SARS-CoV-2 and human rhinovirus are distinguished perfectly from the others, while human coronavirus slightly overlaps with human rhinovirus. This overlap does not necessarily reduce the identification capability for the viruses since a number of spectra (100 or more) are measured per sample and a virus classification score is given based on all of the measured spectra. Since each virus can be identified with 94.1% or 100% recall rates, we can essentially identify perfectly all three viruses at the concentrations and Raman measurement conditions explained above.

### Machine learning classifier for clinical tests

36 COVID-19 PCR positive and 33 PCR negative saliva samples were collected and tested with our Raman metasurface sensor to analyze the clinical diagnostic accuracy of our machine learning models. 200 Raman spectra with 1020 spectral points in each within 600-1700 cm^-1^ were measured from each chip with the sandwich assay (primary aptamer-saliva-secondary aptamer). 15 PCR positive and 12 PCR negative samples (5400 spectra) were used for training the model. A variety of preprocessing methods were applied to the data and the classification accuracies were compared. For Raman spectrum preprocessing, spectrum-wise intensity normalization and feature scaling were implemented. To reduce the number of features, linear discriminant analysis (LDA) was implemented. After LDA, a support vector machine (SVM) algorithm with a linear kernel was implemented. Since 200 spectra were collected for each sample, the test result was calculated using the accuracy score. The percentage of the virus-labeled spectra over the entire spectra gave the overall label of the sample. The optimal “C” value is determined as 0.1, and the decision function shape has been chosen to be “One versus One (OVO)”

The samples have different viral loads which are analyzed using PCR’s CT values ^33^ as the ground truth (or gold standard). The classification performance of the algorithm is related to the variety of CT values in the training and test datasets. **Fig. S6** shows the receiver operating characteristic curves for clinical classification. Here, the different curves correspond to different clinical training sets with various CT distributions and shuffling. Random shuffling of the samples causes the unbalanced distribution of the CT values, which may slightly reduce the classification accuracy (**Fig S6**). To test an extreme case, one balanced and one unbalanced training sets were assembled manually and their test accuracies have been compared. The histograms of these two sets are given in **Fig S7**. The network with a balanced train set has specificity above 90% (**Fig. S7b**). If the network is trained with an unbalanced training dataset (**Fig. S7a**), the specificity results deteriorate significantly (< 50%). Therefore, a balanced training set must be used for improved test accuracy and hence diagnostic sensitivity and specificity.

Our results indicate that our machine learning model can classify the presence of SARS-CoV-2 with 95.2% sensitivity and 92.2% specificity **(Fig. 4b)** when trained with a balanced training dataset. Further extension of the dataset with new clinical samples should improve the robustness of the machine learning model.

### Variant classifier details

3 different variants of SARS-Cov-2 were used to analyze the chip’s and the machine learning models’ ability to distinguish the variants **(Fig. 4c-d**). Specifically, Alpha (B.1.1.7), Beta (B.1.351), and wild-type variants have been used in our chips to measure their Raman spectra. A total of 3225 Raman spectra with 1020 spectrum points in each spectrum were measured in total. There are 1000 spectra from the Alpha variant, 1000 spectra from the Beta variant, and 1225 spectra from the wild-type SARS-CoV-2. The variants were clustered using the t-SNE model ^46^ with the learning rate of 10, component number of 2, perplexity of 30, and the number of iterations of 5000.

The clustering performance of the variant measurements proves Raman spectroscopy as a method that can be used for variant discrimination. In addition to the clustering methods, the classification of the variants was also carried out with the help of supervised machine learning algorithms. After applying spectrum-wise normalization, feature scaling, and principal component analysis (cumulative explained variance of 95% of the data with 474 components), the variants were classified using an SVM algorithm. The optimal C value is determined to be 0.1, gamma is 0.1, the kernel is linear, and the decision function shape has been chosen to be one-versus-one (OVO). 2580 spectra were used for training the network, 645 spectra were reserved for tests (75%:25% training/test dataset ratio). The classification accuracy of test spectra is 98.9%. The sensitivity and the specificity for the alpha variant are 98.4% and 98.9%, for the beta variant are 98.9% and 97.5%, and for the wild-type are 99.2% and 100%, respectively.

## Acknowledgments

The authors gratefully acknowledge the use of the services and facilities of the Koç University Research Center for Translational Medicine (KUTTAM), funded by the Presidency of Turkey, Presidency of Strategy and Budget. The content is solely the responsibility of the authors and does not necessarily represent the official views of the Presidency of Strategy and Budget.

Koç University-İşbank Center for Infectious Diseases (KUISCID) is also gratefully acknowledged for providing the service and facility utilization. Tayfun Barlas and Dr. Gülen Esken from KUISCID are acknowledged for the cells and SARS-CoV-2 isolation and inactivation, and technical support.

Funding support by the Turkish Institutes of Health (TÜSEB) with Project No. 7118-8798 is gratefully acknowledged.

The nanofabrication and characterization infrastructure and services provided by the Sabancı University Nanotechnology Research and Applications Center (SUNUM) are gratefully acknowledged.

BioRender.com is acknowledged for the visual design of Figure 1.

## Funding

Funding support from Turkish Institutes of Health grant no. 7118-8798, Turkish National Academy of Sciences GEBIP Award 2019 (MCO) and The Scientific and Technological Research Council of Turkey grant 119S362 (İS, MCO) are gratefully acknowledged.

## Author contributions

MCO, MI, IS, OE, FC, HT, and BB designed the sensor and articulated the concept.

Data curation: HT, BB, MCO

Formal analysis: BB, HT, MI, MCO, FC

Funding acquisition: MCO, MI, IS, FC, OE, HT, BB

Investigation: HT, BB, CY, SC, MI, MCO

Methodology: MCO, BB, HT, MI, FC, OD, CY, SC, MO

Project administration: MCO, IS

Resources: IS

Software: BB, NB trained and tested the machine learning models.

Supervision: MCO

Validation: MI, FC, OD, BB, HT, MCO

Visualization: MCO, HT, BB, MI

Writing – original draft: MCO, HT, BB, MI, FC, OD

Writing – review & editing: All authors reviewed and commented on the manuscript.

## Competing interests

The authors declare no competing financial interests.

## Data and materials availability

All data used in this study are available from the corresponding author upon request. Institutional Review Board (IRB) approval 2020.112.IRB1.023 restricts the authors from disclosure of patient information or saliva samples. All scripts of this study are available under https://github.com/onbasli/KU-SARS-CoV-2-biosensor/ project GitHub repository.

## Supplementary Materials

Supplementary Information

Figs. S1 to S6

References (1–14)

## Notes

### Competing Interest Statement

The authors have declared no competing interest.

### Clinical Trial

The study has not been registered yet on clinicaltrials.gov, but we registered our clinical trial with Turkish Ministry of Health form number “Mehmet Cengiz Onbaşlı-2020-05-12T06_43_14” under COVID-19 Scientific Research Platform (“Bilimsel Araştırma Platformu”, https://bilimselarastirma.saglik.gov.tr/

)

### Author Declarations

We received Koc University Institutional Review Board (IRB) approval number 2020.112.IRB1.023, which covers all steps of sample collection, informed consent of the volunteers who donated nasopharyngeal and saliva samples at Koc University Hospital's Koc University Clinical Trial Unit.

