## Supplementary Information for "Machine learning detects SARS-CoV-2 and variants rapidly on DNA aptamer metasurfaces"

**Supplementary Materials for “Machine learning detects SARS-CoV-2 and variants rapidly on DNA aptamer metasurfaces”**

**Fluorescent secondary aptamer design and primary aptamer design with a thiol linker**

Nucleic acid aptamers have become an alternative to antibodies in diagnostics due to their stability, low cost, reusability, easy chemical modification, and reproducible synthesis ^1-6^. As highly specific bio-reagents, they offer fast and accurate identification of disease-causing agents or the resulting cues produced by the infected body.

DNA aptamers have been obtained against SARS-CoV-2 spike protein in a recent study ^7^. We used one of these DNA aptamers namely 4RBD-4C in our studies (Figure S1) and verified its ability to interact with SARS-CoV-2 particles (Figure S1). It allowed sensitive detection of COVID-19 infection in an aptasensor sandwich assay, in which the primary aptamer was immobilized on the SERS surface and the secondary was labeled with Cy5.5 as a Raman reporter molecule.

Previous studies demonstrated that Raman spectroscopy can be used for DNA aptamer-based biosensing ^8-12^. This novel aptasensor has the potential for the rapid and sensitive diagnosis of SARS-CoV-2.

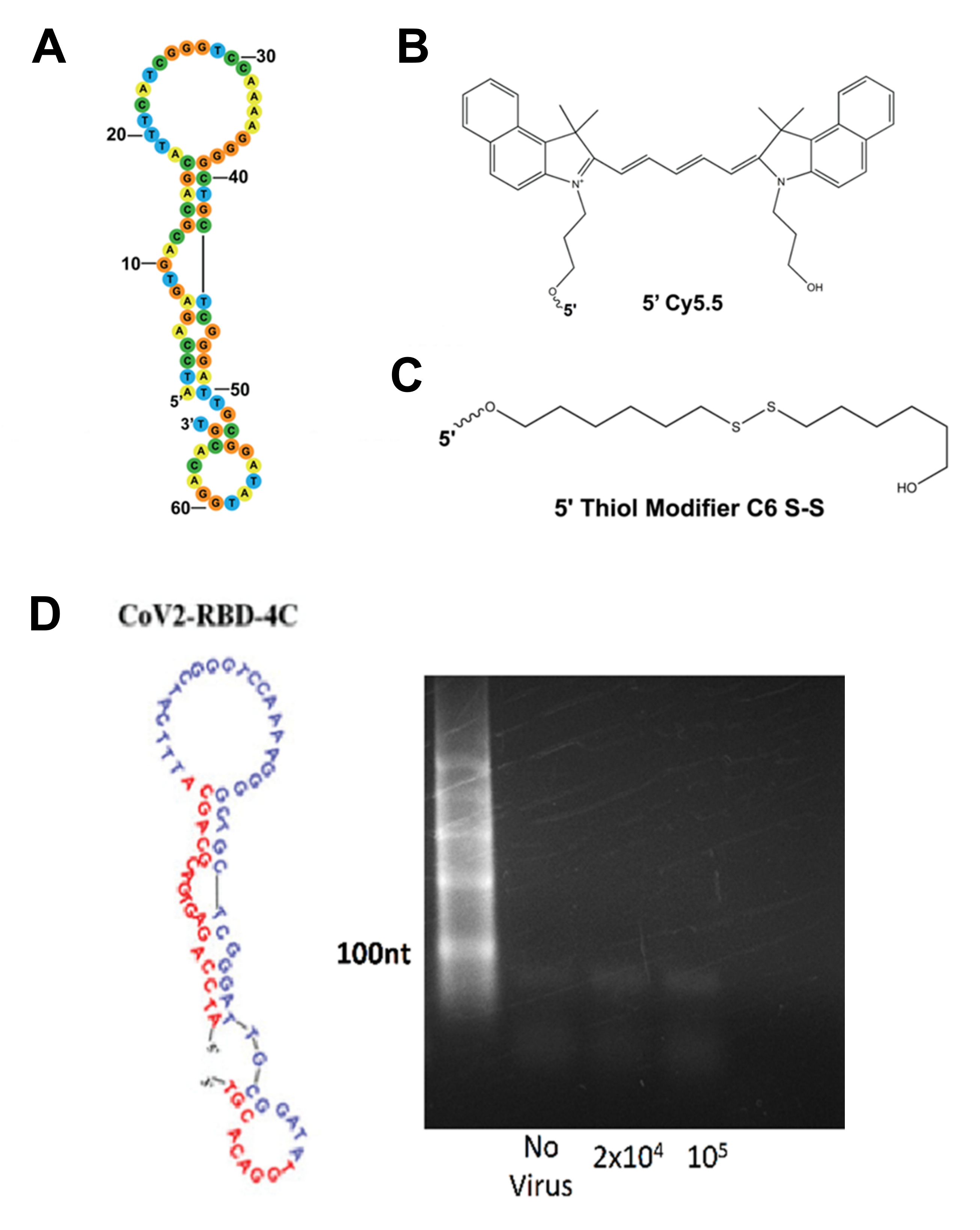

Fig. S1. Primary DNA aptamer design with a thiol linker and the secondary DNA aptamer with a fluorescent marker. **a,** The aptamer structure and its sequence for binding on the virus. This aptamer has been used for primary and secondary aptamers. **b,** Cy5.5 fluorescent molecule and its structure. **c,** The thiol linker layer for binding the primary aptamer on the gold metasurface. The primary aptamer has been bound to the thiol modifier C6 shown in **Fig. S1c** to functionalize the gold metasurface. The secondary aptamer has been modified with the fluorescent marker molecule Cy5.5, which absorbs around the red (600-700 nm) and emits in the near-infrared wavelengths (680-800 nm). The Raman spectra were measured within 600-1700 cm^-1^ (Raman shifts) or 658-710 nm wavelengths. **d**. The structure of the CoV-2-RBD-4C DNA aptamer used from reference ^7^ is shown and the confirmation of SARS-CoV-2 binding of the aptamer. Thiol group (-SH) was used to modify the aptamer as the primary aptamer. Cy5.5 fluorescent marker was attached to the same aptamer to be used as the secondary aptamer in the SERS metasurface biosensor.

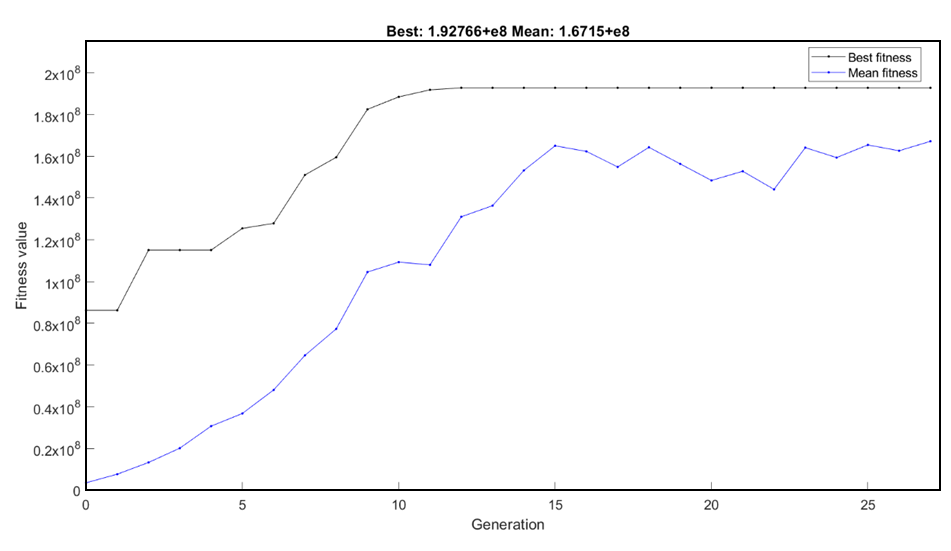

Fig. S2. Genetic algorithm model fitness parameter evolution as a function of generation. Our genetic algorithm and finite-difference time-domain electromagnetic modeling iteratively refined the geometries to yield the highest fitness value, which is defined as the average electric field enhancement factor (η = |E|^4^/|E_0_|^4^, where E_0_ is the incident electric field.). As further mutations and crossover in 15 generations did not change the fitness value, the genetic algorithm stopped geometric refinement.

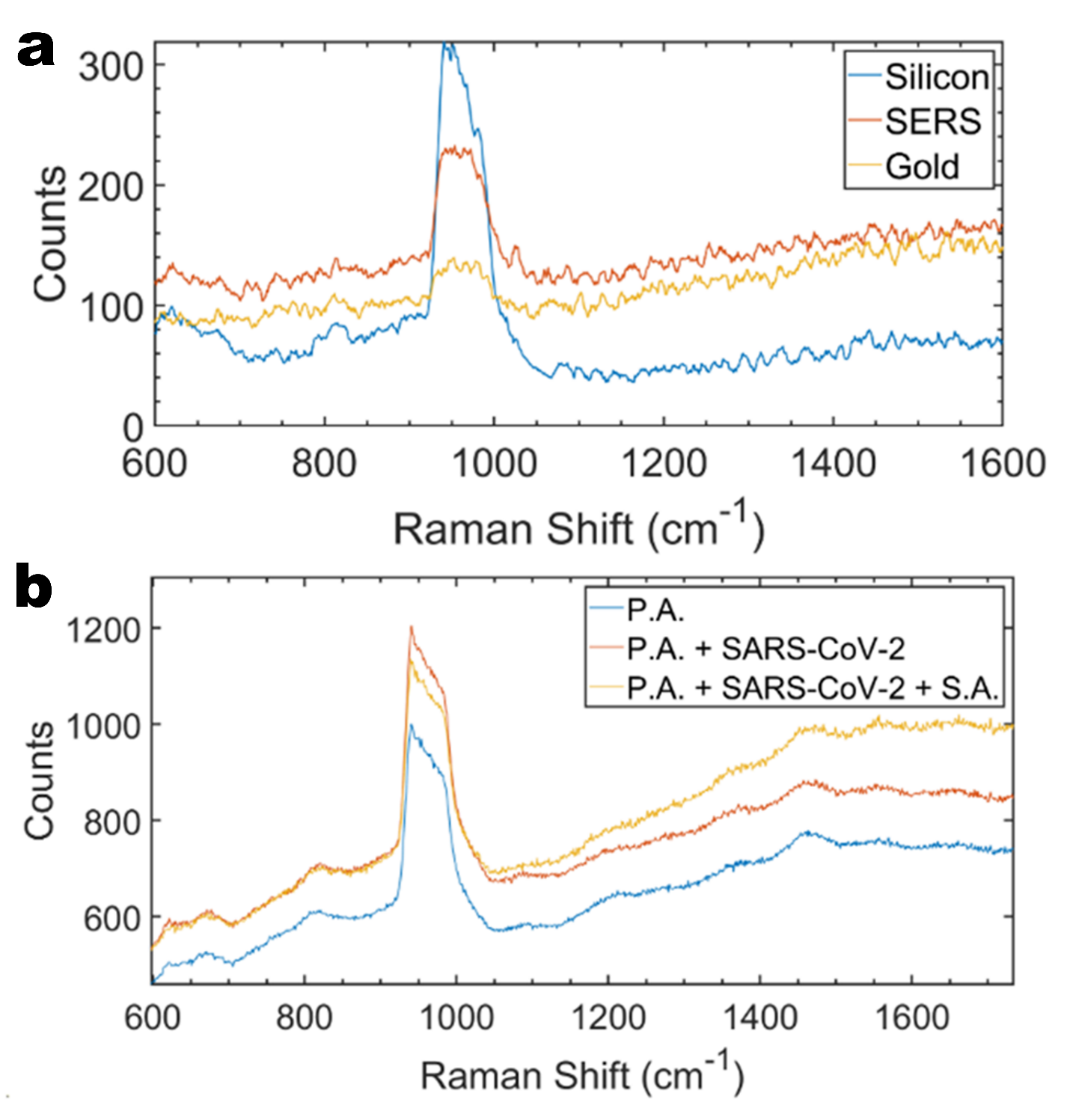
Fig. S3. Raman spectral baseline measurements (Si, planar gold on Si, SERS, P.A. on SERS, sandwich assay on SERS). **a,** Raman spectra of bare silicon, planar gold-coated silicon, and SERS metasurface without any aptamers are shown. The main peak centered at 970 cm^-1^ (within 920-1020 cm^-1^) originates from the silicon substrate’s strong transverse optical phonon line ^13,14^. The SERS metasurface on silicon leads to broadband enhancement of the Raman signal. **b,** The Raman spectra of the primary aptamer (P.A.), primary aptamer bound with SARS-CoV-2, and the sandwich assay of primary aptamer bound to the virus which is bound to the fluorescent secondary aptamer (S.A.). The metasurface enhances the fluorescent broadband Raman slope, multiple peaks as well as the 1575 cm^-1^ Raman peak originating from the virus (**Fig. 3a** in the main manuscript).

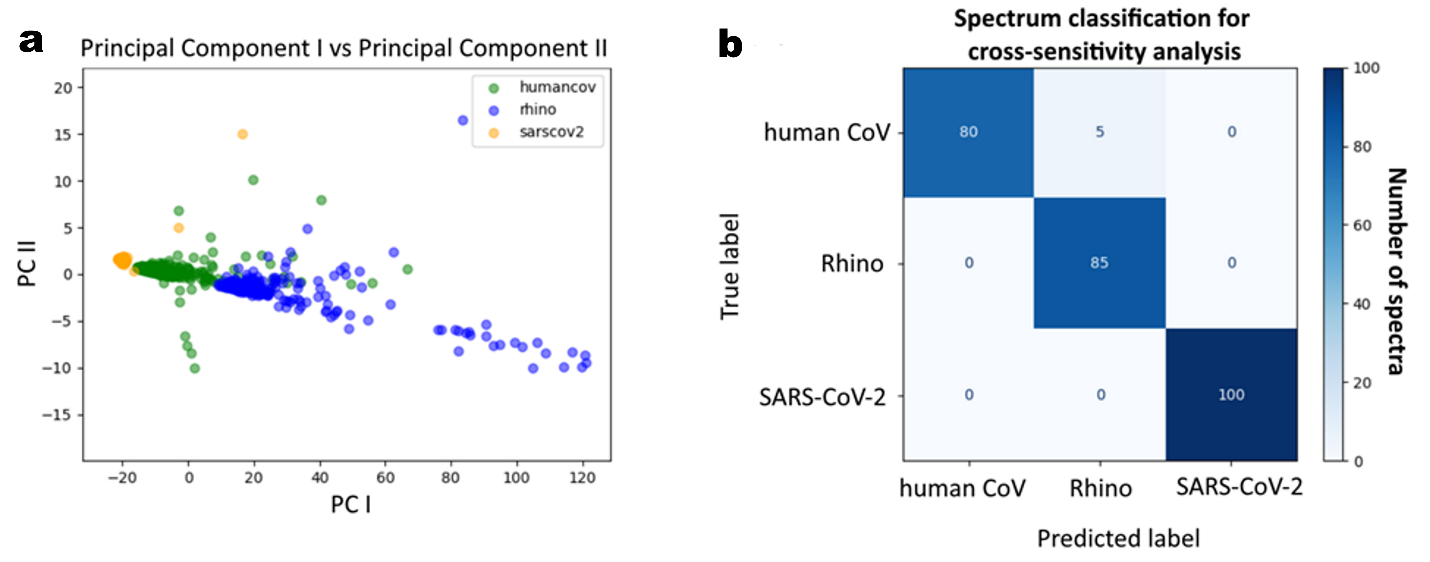
Fig. S4. Machine learning-based identification of SARS-CoV-2, human rhinovirus, and human coronavirus from cross-sensitivity tests. **a,** The principal component analysis (PCA) shows a clear distinction of the Raman spectra for human rhinovirus, human coronavirus, and SARS-CoV-2 (10^8^ pfu/ml) sandwich assay on our metasurface chip. These spectra were measured under the same Raman measurement parameters as for the clinical samples. The axes are the two principal components that are used for identifying the virus type. **b,** The confusion matrix shows a near-perfect (99.7% cumulative variance) accuracy in multiclass classification results and a successful distinction of these three different viruses. The fact that this distinction could be made using only two principal components shows the power of Raman and PCA in identifying if there is any cross-sensitivity of the metasurface to other viruses.

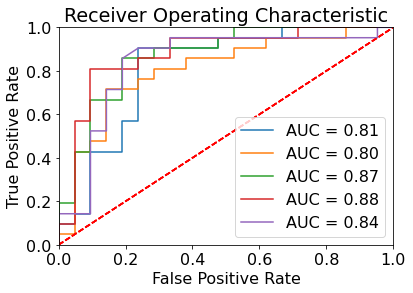

Fig. S5. ROC curves for the clinical test results for different shuffling conditions. Clinical saliva samples were tested, and their receiver operating characteristics are obtained for a variety of shuffling conditions where more unbalanced training datasets of CT (cycle threshold) values for the saliva samples lead to lower areas under the curve (AUC). For best classification accuracy, a training dataset that consists of balanced CT value ranges is required. In the optimal training set, our machine learning model yields 95.2% sensitivity and 95.2% specificity on 42 test samples **(Fig. 4b).**

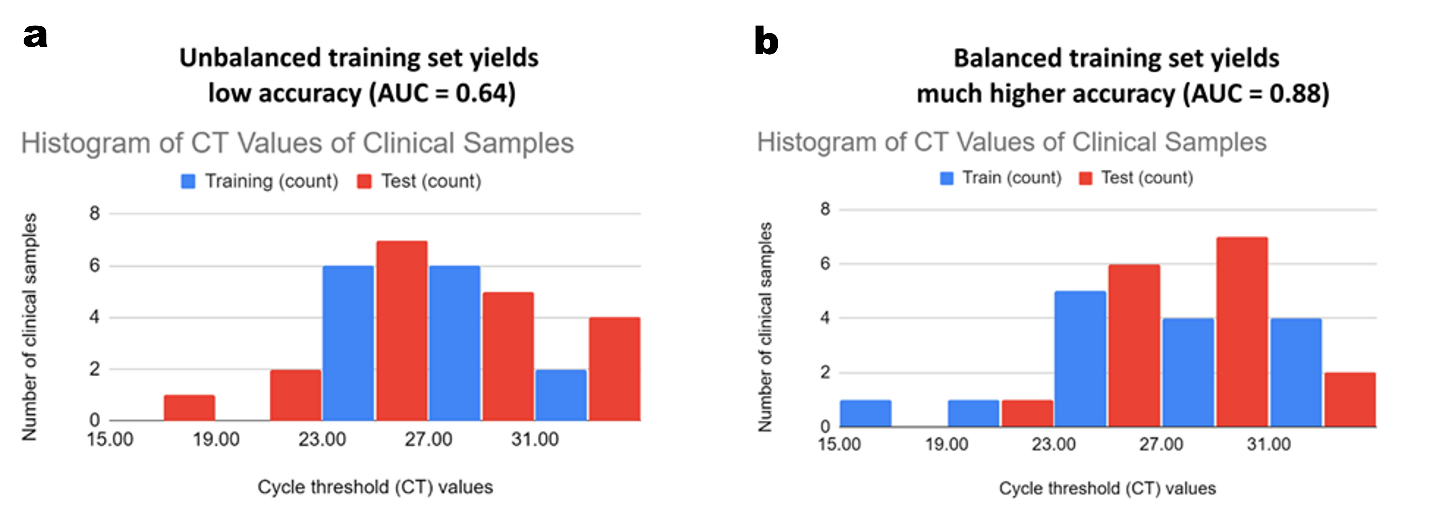
Fig. S6. Effect of CT distribution in the training set on the machine learning model accuracy. Two training sets were manually assembled for evaluating the effect of an unbalanced distribution of training set CT values on the classification accuracy and areas under the curve (AUC). **a,** The unbalanced dataset that does not include clinical samples with high viral loads (CT < 23) and yields a low AUC. **b,** The balanced training dataset that includes samples with high viral loads yield higher classification accuracies. For the highest classification accuracy, a training dataset that consists of a balanced CT value range is required.
